# Mammography Access, Urbanicity, and Late-Stage Breast Cancer Burden Across Texas: A Bayesian Spatial Analysis

**DOI:** 10.64898/2026.07.11.26357817

**Authors:** Yue Zhang, Jilin Tian, Gayla M. Ferguson, Bryan Colby Griffin, Jason H. Windett, Jingjing Gao

## Abstract

**Background:** Geographic disparities in access to preventive healthcare services remain an important contributor to breast cancer inequities in the United States. Mammography screening plays a critical role in early detection and improved survival; however, screening infrastructure and healthcare accessibility remain unevenly distributed across many regions, particularly within large and socioeconomically diverse states such as Texas. Understanding the spatial relationships among mammography access, urbanicity, socioeconomic vulnerability, and late-stage breast cancer burden is important for developing geographically targeted public health interventions.

**Methods:** This study integrated multiple county-level datasets, including mammography facility locations from the Texas Cancer Information database, late-stage breast cancer burden data from the National Cancer Institute, and socioeconomic indicators from census-derived datasets and the Centers for Disease Control and Prevention Social Vulnerability Index. Geographic information systems (GIS), spatial autocorrelation analyses, and Bayesian spatial epidemiologic methods were used to evaluate geographic patterns across Texas counties from 2018 to 2022. Global and local Moran’s I statistics were calculated to assess spatial clustering patterns. Bayesian spatial Poisson conditional autoregressive (CAR) regression models were subsequently estimated to examine associations between mammography center density, population density, female socioeconomic characteristics, and late-stage breast cancer burden while accounting for residual spatial dependence.

**Results:** Significant positive spatial autocorrelation was observed for county-level late-stage breast cancer burden across Texas counties. Mammography facilities were heavily concentrated within major metropolitan regions, while many rural counties demonstrated comparatively limited screening infrastructure availability. Bayesian spatial regression analyses demonstrated that log-transformed population density was significantly inversely associated with the burden of late-stage breast cancer (β = -0.136, 95% CrI [-0.175, - 0.103]), indicating that less densely populated counties experienced greater burden than more urbanized counties. Mammography center density showed a borderline inverse association with late-stage burden (β = -0.008, 95% CrI [-0.017, 0.002]), suggesting that greater availability of screening infrastructure may contribute to reduced burden. Persistent residual spatial dependence remained across counties (ρ = 0.472, 95% CrI [0.038, 0.919]), indicating ongoing geographic clustering beyond measured explanatory variables.

**Conclusions:** Substantial geographic disparities in late-stage breast cancer burden, mammography access, and socioeconomic vulnerability exist across Texas counties. The findings suggest that urbanicity and screening infrastructure availability play important roles in shaping geographic inequities in breast cancer outcomes. Public health interventions should move beyond increasing facility availability alone and instead incorporate geographically targeted strategies that address rural healthcare access limitations, healthcare infrastructure disparities, and broader structural barriers to preventive screening services.

## Background

Breast cancer is the most frequently diagnosed cancer among women in the United States. It remains a leading cause of cancer-related mortality worldwide, with over 2.3 million new cases reported in 2022, and incidence is expected to exceed 3 million cases annually by 2040 [1]. According to the American Cancer Society’s 2026 estimates, approximately 321,910 new cases of breast cancer were diagnosed in women, as well as an estimated 42,140 deaths [2]. In the United States, breast cancer incidence has continued a steady upward trend, rising by 1% annually between 2013 and 2022. In contrast, the overall breast cancer death rate declined by 44% from 1989 to 2023. This substantial reduction in mortality is mainly attributed to widespread screening mammography initiatives that enable early detection, along with continuous advancements in targeted treatments [2, 3].

Breast cancer is strongly associated with age, sex, and genetic susceptibility, with older females experiencing substantially greater risk because of cumulative hormonal exposure, age-related cellular changes, and inherited genetic predisposition factors such as BRCA1 and BRCA2 mutations [2, 4–8]. Besides age and sex, genetic predispositions [9], family history of breast cancer [1], and reproductive and hormonal history [4] are all non-modifiable factors that would drastically increase risk. Modifiable lifestyle factors, including obesity, alcohol consumption, and smoking, have all been consistently linked to higher incidences of the disease [5]. Furthermore, significant racial and socioeconomic disparities remain in breast cancer outcomes. For example, although Black women have a slightly lower breast cancer incidence rate than White women, they experience substantially higher mortality and lower survival rates across nearly all disease subtypes and stages [3]. While biological and behavioral factors dictate baseline risk, systemic factors such as poverty, lack of insurance, and geographic barriers determine the stage at which the disease is diagnosed [10, 11]. These disparities highlight the critical role of social determinants of health, including socioeconomic status (SES) and access to healthcare, in lowering breast cancer incidence rates.

Previous research has demonstrated that population-level mammography screening programs are associated with meaningful reductions in breast cancer mortality, with several large studies estimating mortality reductions of approximately 20% among screened populations [12]. Moreover, participation in regular screening programs is directly associated with a substantial reduction in the incidence of advanced and fatal breast cancers [13]. Despite these clear benefits, it also has some limitations and trade-offs. Specifically, widespread screening increases the risk of false-positive results, as well as overdiagnosis and subsequent overtreatment [12, 14]. These limitations underscore the need to examine whether people have timely, equitable access to high-quality screening.

Geographic accessibility to healthcare facilities is a crucial determinant of screening use and stage at diagnosis. Measures of access in the literature include facility capacity, density, distance, and travel time, while findings have been mixed. Systematic reviews have demonstrated that women living in geographically isolated or underserved areas, such as rural counties or low-income urban neighborhoods, are significantly less likely to receive regular mammograms [11, 15]. Thus, limited spatial access is strongly correlated with a higher incidence of late-stage breast cancer diagnosis. However, several studies indicate that the impact of geographic access on breast cancer screening is heterogeneous and modified by the community’s SES. Recent literature suggests that the combined effects of SES and geographic access should be studied simultaneously to fully capture the spatial disparities in late-stage diagnoses [10, 11]. A spatial mismatch between high-risk populations with disadvantages of SES and the distribution of mammography clinics can lead to severe delays in diagnosis.

Texas is a highly complex and unique landscape for studying spatial healthcare disparities. As the second-largest state by both area and population, Texas encompasses sprawling metropolitan centers and sparsely populated rural regions. In 2026, it is estimated that 24,270 women in Texas will be newly diagnosed with breast cancer, and about 3,300 individuals will die from the disease [2]. Beyond its vast geographic size, Texas has a diverse demographic and socioeconomic profile, including a large proportion of residents living below the poverty line and the highest rate of uninsured individuals in the country [16–22]. These characteristics of the state would create barriers to preventative care [16, 23–25]. As a result, understanding how the distribution of mammography clinics interacts with these localized demographic factors is essential for addressing the state’s late-stage breast cancer burden.

Identifying geographic inequities in mammography infrastructure is directly relevant to cancer prevention and control because delayed or inconsistent access to screening may contribute to later-stage diagnosis, higher treatment burden, and worse survival outcomes. Spatial analyses can help cancer-control programs identify counties where screening infrastructure, rurality, and socioeconomic vulnerability overlap, allowing public health agencies to prioritize mobile mammography, transportation support, patient navigation, and targeted outreach.

We aim to evaluate the spatial relationship between geographic access to mammography clinics and the incidence rate of late-stage breast cancer while accounting for socioeconomic status across Texas counties. Specifically, this study seeks to map the spatial distribution of mammography clinics and late-stage breast cancer rates and to identify spatial autocorrelation and statistically significant geographic clusters of these variables. We hypothesize that counties with lower mammography center availability, lower population density, and greater socioeconomic vulnerability will experience significantly greater late-stage breast cancer burden. We further hypothesize that late-stage breast cancer burden will demonstrate significant spatial clustering across Texas counties, reflecting underlying geographic disparities in healthcare infrastructure and preventive screening accessibility.

## Methods

### Study Area

Texas is located in the South-Central region of the United States, bounded by the Gulf of Mexico, Mexico, New Mexico, Oklahoma, Arkansas, and Louisiana. It encompasses an area of approximately 268, 596 square miles, featuring a highly diverse climate and terrain that ranges from eastern coastal plains and pine forests to central hills and western desert mountains. As of 2025, the population was estimated to be 31,709,82, and this population is distributed across 254 counties (U.S. Census Bureau, 2025).

### Data Sources

This study integrated multiple publicly available county-level datasets to examine the spatial relationships among mammography access, socioeconomic vulnerability, urbanicity, and late-stage breast cancer burden across Texas counties from 2018 to 2022. The primary geographic unit of analysis was the county level, consistent with the availability of statewide cancer surveillance and socioeconomic data. Mammography facility locations were obtained from the Texas Cancer Information database (2025), which provides geographic information on certified mammography screening centers across Texas. Facility addresses were geocoded and spatially linked to county boundaries to calculate mammography center density measures for each county. County-level late-stage breast cancer burden data were obtained from the National Cancer Institute’s State Cancer Profiles database, which provides population-based cancer surveillance statistics across Texas counties. Socioeconomic and demographic characteristics, including female population composition, unemployment, insurance status, and educational vulnerability indicators, were derived from the National Historical Geographic Information System (NHGIS) and Centers for Disease Control and Prevention Social Vulnerability Index datasets. County boundary shapefiles used for spatial mapping and geographic analyses were obtained from the United States Census Bureau TIGER/Line database. All datasets were harmonized and merged using county-level geographic identifiers to construct a unified spatial analytic dataset for Texas counties. Geographic information system (GIS) methods were subsequently used to integrate spatial healthcare infrastructure data with population-level socioeconomic and cancer burden indicators.

### Analysis Plan

Spatial analyses were conducted using ESRI ArcGIS Pro, GeoDa, and R statistical software to examine the geographic relationship between mammography access, urbanicity, socioeconomic vulnerability, and late-stage breast cancer burden across Texas counties from 2018 to 2022. Mammography clinic addresses were geocoded to generate spatial point data, allowing for the calculation of county-level mammography center density per 100,000 female population. County-level late-stage breast cancer burden, population density, and female socioeconomic indicators were mapped to visualize geographic distributions and identify potential spatial disparities across Texas.

The dependent variable for the analysis was a county-level late-stage breast cancer burden. Independent variables included mammography center density, log-transformed population density, female uninsured percentage, female unemployment percentage, and female population composition indicators. Educational vulnerability was additionally assessed using the percentage of adults without a high school diploma derived from the CDC Social Vulnerability Index (SVI).

To evaluate spatial dependence and clustering patterns, several spatial statistical techniques were employed. Univariate Global Moran’s I statistics were calculated to assess overall spatial autocorrelation for late-stage breast cancer burden and primary explanatory variables across Texas counties. Univariate Local Indicators of Spatial Association (LISA) analyses were subsequently conducted to identify statistically significant local hotspots (High-High clusters), coldspots (Low-Low clusters), and spatial outliers (High-Low and Low-High clusters).

Bivariate Global Moran’s I statistics were then calculated to evaluate the overall spatial relationships between late-stage breast cancer burden and key explanatory variables, including mammography center density and female uninsured percentage. Bivariate LISA analyses were additionally performed to identify localized spatial relationships between late-stage breast cancer burden and healthcare access indicators across Texas counties.

Because significant spatial autocorrelation was identified, Bayesian spatial Poisson conditional autoregressive (CAR) regression models were subsequently estimated to account for residual geographic dependence across counties [26–30]. County-level late-stage breast cancer counts served as the outcome variable, while the natural logarithm of the female population was included as an offset term to account for differences in county population size. Model performance was evaluated using Deviance Information Criterion (DIC), Widely Applicable Information Criterion (WAIC), and Log Marginal Predictive Likelihood (LMPL), with lower DIC and WAIC values indicating improved model fit.

## Results

### Descriptive Spatial Characteristics of Late-Stage Breast Cancer Burden Across Texas Counties

Table 1a presents descriptive statistics for county-level variables included in the spatial analysis across Texas counties. The mean county-level late-stage breast cancer count was 25.67 cases (SD = 88.26), although substantial variability was observed across counties, with counts ranging from 2 to 1,020 cases. Mammography center density averaged 4.46 centers per 100,000 female population (SD = 7.42), indicating considerable geographic variation in screening infrastructure availability across the state.

Log-transformed population density demonstrated substantial heterogeneity across Texas counties (mean = 3.21, SD = 1.68), reflecting the coexistence of highly urbanized metropolitan counties and sparsely populated rural counties. The average female population across counties was 57,553.95 individuals, although county populations varied considerably because of the large geographic and demographic diversity of Texas.

Female unemployment percentage averaged 4.3%, while female uninsured percentage averaged 16.6%, suggesting notable socioeconomic variation across counties. Overall, the descriptive findings demonstrated substantial geographic heterogeneity in healthcare infrastructure, population distribution, and socioeconomic characteristics across Texas counties, supporting the appropriateness of spatial epidemiologic modeling approaches.

**Table 1a.** Descriptive Statistics of County-Level Variables Included in the Spatial Analysis Across Texas Counties.

| **Variable** | **Mean** | **SD** | **Median** | **Min** | **Max** |
| --- | --- | --- | --- | --- | --- |
| Late-stage breast cancer count | 25.669 | 88.258 | 5.000 | 2.000 | 1020.000 |
| Mammography centers per 100,000 female population | 4.459 | 7.416 | 1.964 | 0.000 | 59.032 |
| Log-transformed population density | 3.213 | 1.679 | 3.114 | 0.077 | 8.001 |
| Female population | 57,553.953 | 206,128.668 | 9,111.000 | 45.000 | 2,366,762.000 |
| Female population percentage (%) | 0.488 | 0.035 | 0.499 | 0.352 | 0.595 |
| Female unemployment percentage (%) | 0.043 | 0.029 | 0.040 | 0.000 | 0.186 |
| Female uninsured percentage (%) | 0.166 | 0.055 | 0.163 | 0.000 | 0.340 |

**Note:** Descriptive statistics were calculated across Texas counties included in the spatial epidemiologic analysis. Population density was log-transformed because of substantial right skewness in county-level population distribution. Mammography center density was calculated as the number of mammography centers per 100,000 female population.

**Figure 1:**
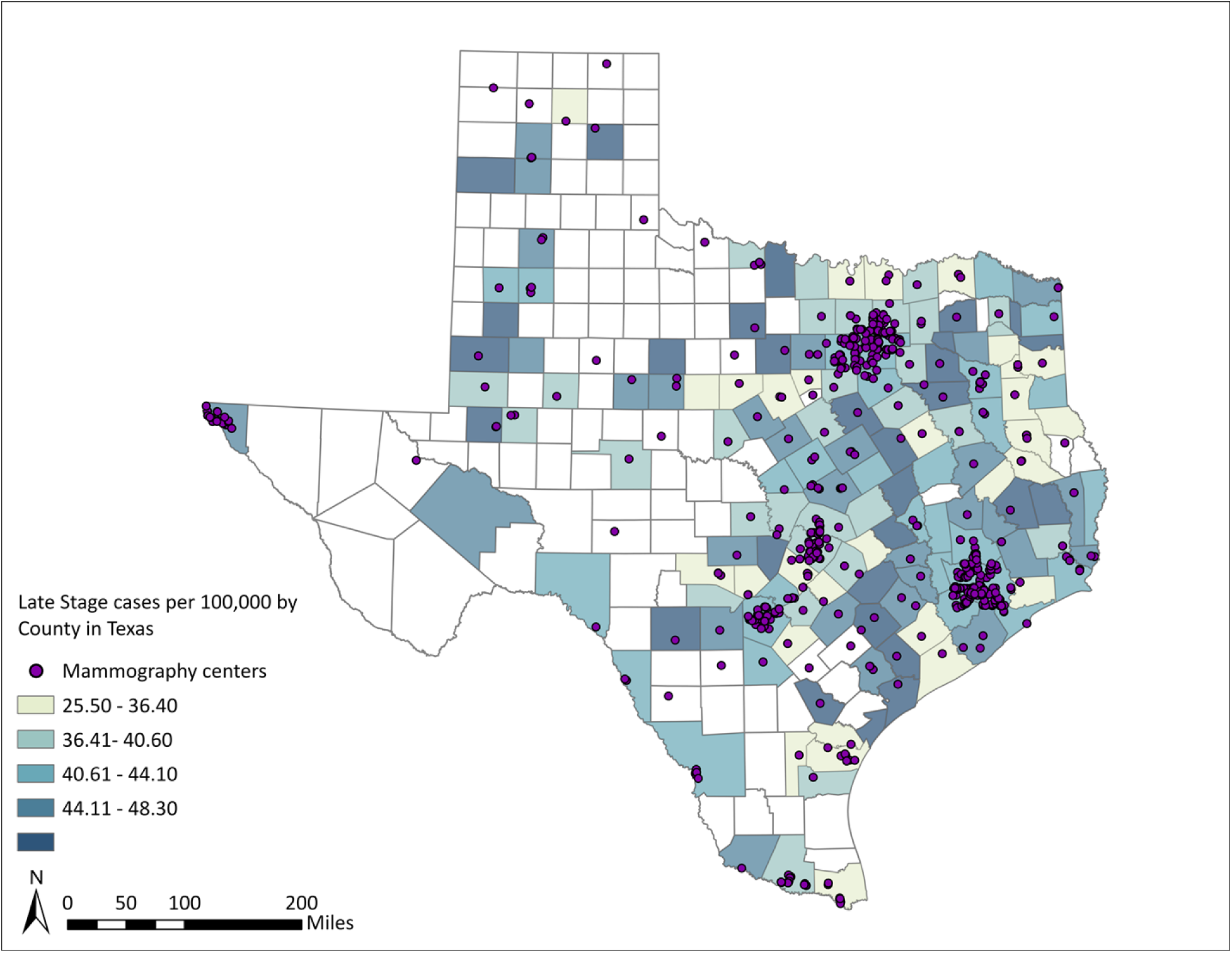
Spatial Distribution of Incidence Rates for Texas by County

Figure 1 presents the spatial distribution of county-level late-stage breast cancer incidence rates across Texas counties. Elevated late-stage breast cancer burden was concentrated primarily within portions of East Texas, Central Texas, and selected southeastern counties. Several clusters of counties with comparatively higher incidence rates were observed adjacent to neighboring counties with similarly elevated burdens, suggesting potential geographic clustering. In contrast, lower incidence rates were more common in portions of West Texas and sparsely populated rural regions. Several counties contained suppressed or unavailable values because of privacy protection procedures associated with low case counts. This figure also illustrates the geographic distribution of mammography service locations across the state. In total, there are 625 mammography centers, with 103 centers located in Harris County, 51 centers in Dallas County, and 42 centers in Bexar County. In contrast, large portions of West Texas, the Panhandle, and several rural counties demonstrated sparse mammography infrastructure availability. These findings suggest substantial geographic disparities in access to breast cancer screening services across Texas counties.

**1**Overall, the visual spatial distributions shown in Figures 1 demonstrated notable geographic heterogeneity in mammography infrastructure, socioeconomic vulnerability, and late-stage breast cancer burden across Texas. The observed clustering patterns further supported the use of formal spatial autocorrelation analyses and Bayesian spatial regression modeling to account for geographic dependence across counties.

### Global and Local Spatial Autocorrelation Analysis of Late-Stage Breast Cancer Burden and Mammography Access

**Table 1b.** Global Moran’s I Results.

|  | Variable | Moran's I value | Pseudo p-value |
| --- | --- | --- | --- |
| **Univariate** | Late-stage incidence rate | 0.312 | < 0.05 |
|  | Number of mammography centers | 0.151 | < 0.05 |
|  | Percentage of Uninsured female | 0.249 | < 0.05 |
| **Bivariate** | X = Number of mammography centers | 0.156 | < 0.05 |
|  | Y = Late-stage incidence rate |  | < 0.05 |
|  | X = Percentage of uninsured females | -0.145 | < 0.05 |
|  | Y = Late-stage incidence rate |  | < 0.05 |

To assess the overall spatial trends across Texas, global Moran’s I statistics were calculated for all primary variables (Table 1b and Figure 4). The univariate global Moran’s I result indicated significant positive spatial autocorrelation across the state for all three variables since all p-values were less than 0.05. Late-stage breast cancer incidence rates showed the strongest positive spatial clustering (Moran’s I = 0.312), followed by the percentage of uninsured females (Moran’s I = 0.249) and the number of mammography centers (Moran’s I = 0.151). Bivariate global Moran’s tests were also conducted to evaluate the statewide spatial relationships between the dependent and independent variables. The relationship between the number of mammography centers and late-stage incidence rates suggested a positive spatial correlation (Moran’s I = 0.156, *p* < 0.05). However, the relationship between the percentage of uninsured females and late-stage incidence rates showed a negative global spatial correlation (Moran’s I = -0.145, *p* < 0.05). While global metrics provide a statewide overview, local analyses are required to discover specific geographic disparities and localized clusters underlying the global patterns.

**Figure 2.**
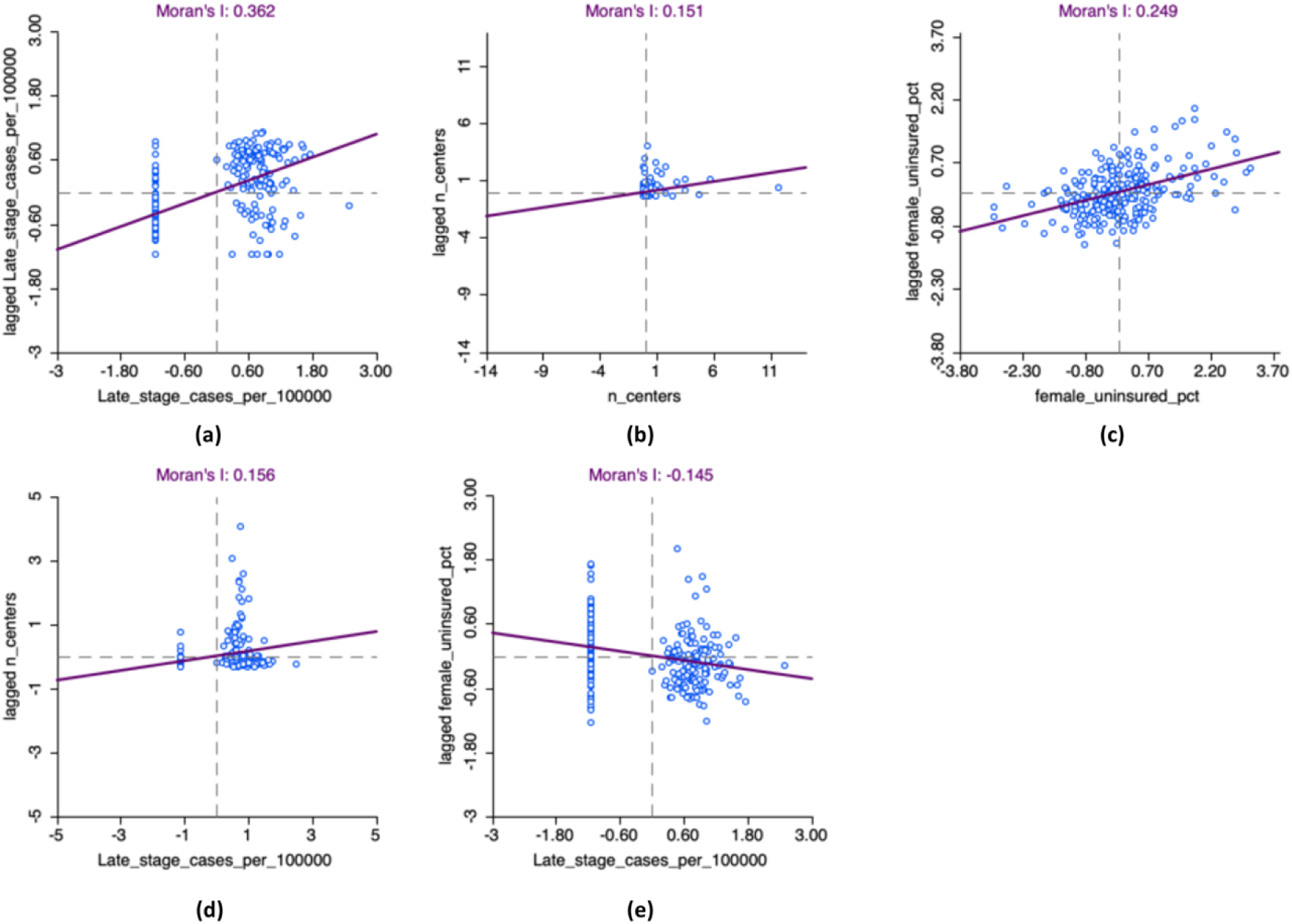
Univariate and Bivariate Global Moran’s I Scatter Plots. (a) Univariate Moran scatter plot for late-stage breast cancer incidence rate. (b) Univariate Moran scatter plot for the number of mammography centers. (c) Univariate Moran scatter plot for the percentage of uninsured females. (d) Bivariate Moran scatter plot illustrating the spatial correlation between late-stage incidence and the number of mammography centers. (e) Bivariate Moran scatter plot illustrating the spatial correlation between late-stage incidence and the percentage of uninsured females.

**Table 1c.**
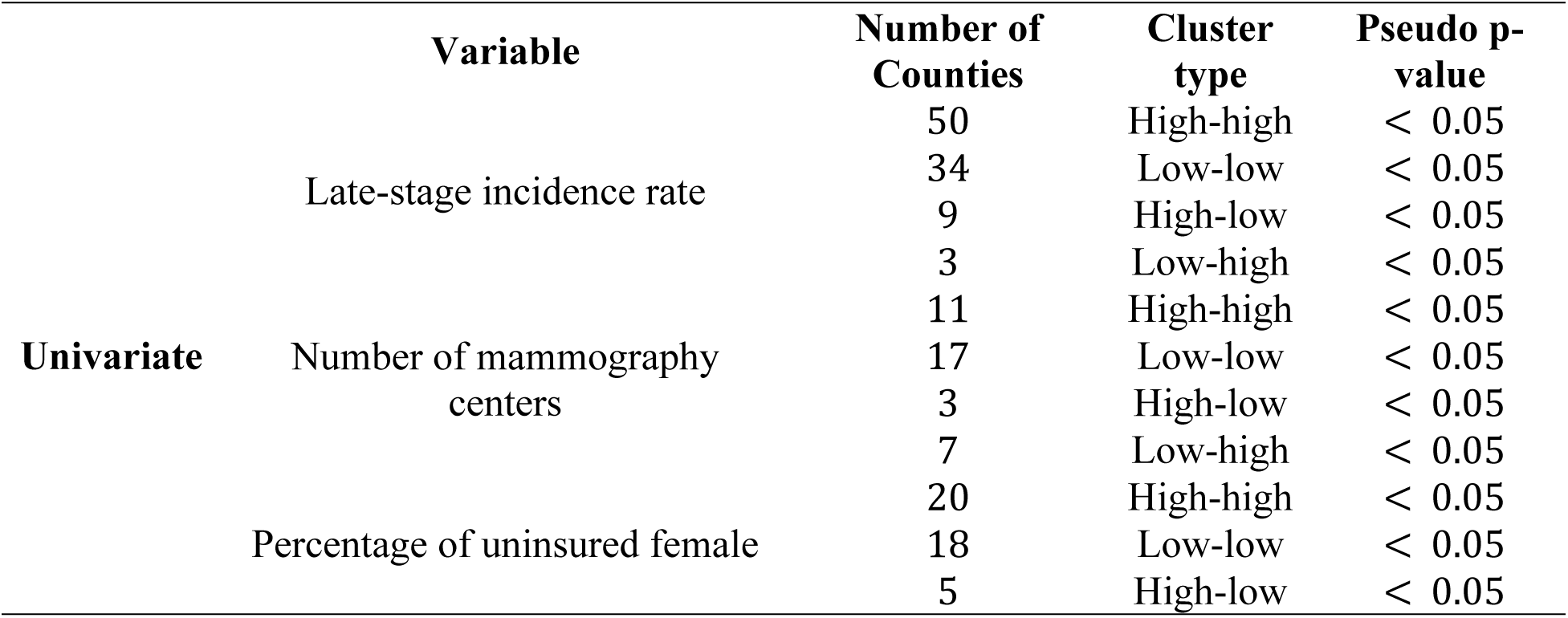
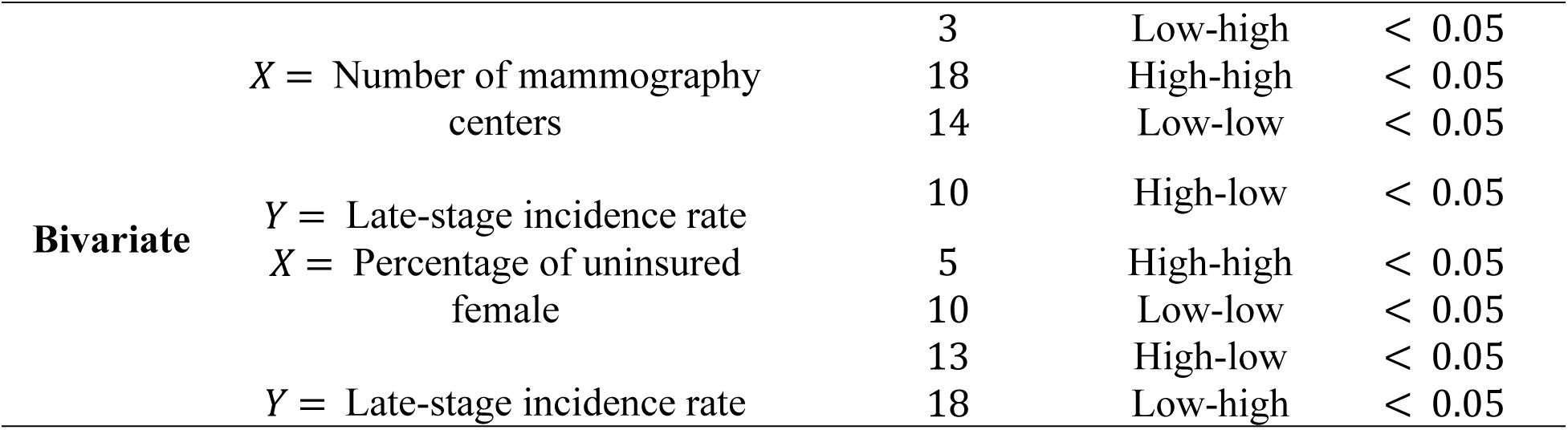
Local Moran’s I Results.

The univariate LISA analysis revealed highly distinct spatial structures for healthcare access, socioeconomic status, and health outcomes across Texas (Table 1b and Figures 5-6). For late-stage incidence rates, the analysis identified 50 counties that were significant High-High clusters, primarily concentrated in East and Central Texas, indicating counties with persistently elevated late-stage breast cancer burden surrounded by neighboring counties with similarly high burden. Many of these counties were characterized by lower population density, reduced mammography infrastructure availability, higher socioeconomic vulnerability, and limited healthcare accessibility, suggesting that structural and geographic barriers may contribute to delayed breast cancer detection within these regions. The geographic concentration of these clusters further indicates that late-stage breast cancer burden in Texas is not randomly distributed, but instead reflects broader regional inequalities in healthcare access and preventive screening resources. Meanwhile, 34 counties were significant Low-Low clusters, predominantly located in the Panhandle and West-central Texas. Significant hotspots for geographic access to screening were identified exclusively around major metropolitan areas, such as the Dallas-Fort Worth, Houston, and San Antonio regions. In addition, cold spots were mainly located in rural areas. Socioeconomic vulnerability showed a stark geographic divide. Hotspots of uninsured population (20 counties) were heavily concentrated along the Texas-Mexico border and portions of the Panhandle. In contrast, 18 counties formed cold spots, representing areas of higher insurance coverage, located primarily in the suburban rings of major cities and in parts of Central Texas.

**Figure 3.**
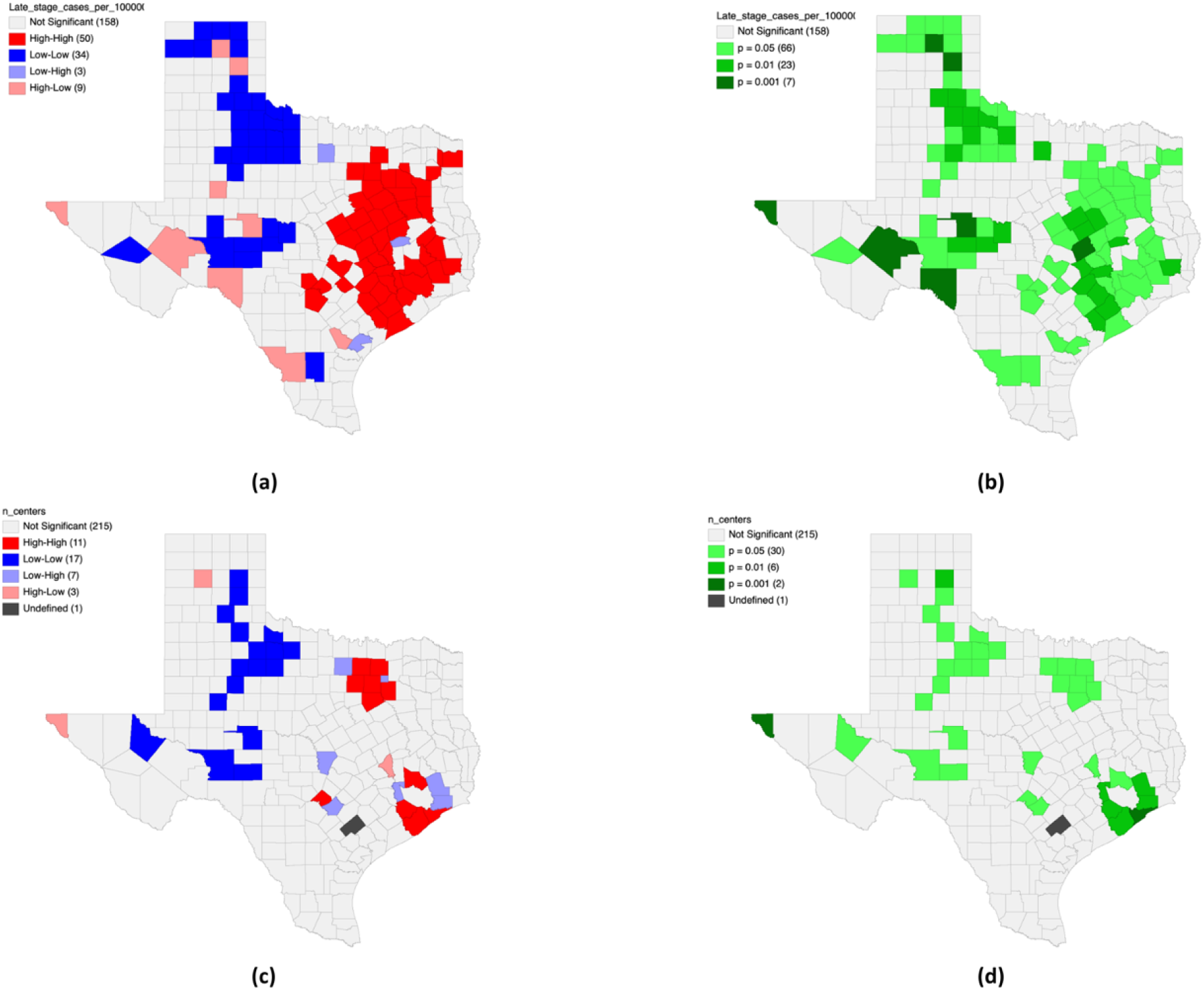
Univariate Local Indicators of Spatial Association (LISA) for Late-Stage Incidence and Mammography Centers. (a) Univariate LISA cluster map for late-stage breast cancer incidence rates per 100,000. (b) Corresponding LISA significance map for late-stage incidence rates. (c) Univariate LISA cluster map for geographic access, measured by the number of mammography centers per county. (d) Corresponding LISA significance map for the number of mammography centers.

Bivariate LISA analyses were utilized to assess how localized geographic access and socioeconomic barriers interact with late-stage cancer outcomes (Table 1b and Figures 6-7). The bivariate analysis for late-stage incidence and neighborhood mammography center availability identified 18 High-High counties, mostly in urban centers, where high clinic density was spatially associated with high late-stage incidence rates. Additionally, there were 14 Low-Low counties located in the Panhandle and West-central Texas, where those counties had low incidence rates and nearby counties also had low mammography center availability. The map also identified significant High-Low outliers, primarily in Central and North Texas, where counties with high late-stage incidence were surrounded by counties with low mammography center availability. The relationship between late-stage incidence and percentage of uninsured female was more complex as the global bivariate Moran’s I was negative, suggesting that counties with high late-stage incidence were not consistently surrounded by counties with high uninsured burdens. This was supported by the bivariate. Although several High-High clusters (5 counties) were present along the southern Texas-Mexico border, the overall pattern was characterized by substantial High-Low and Low-High outliers that distributed across the state.

**Figure 4.**
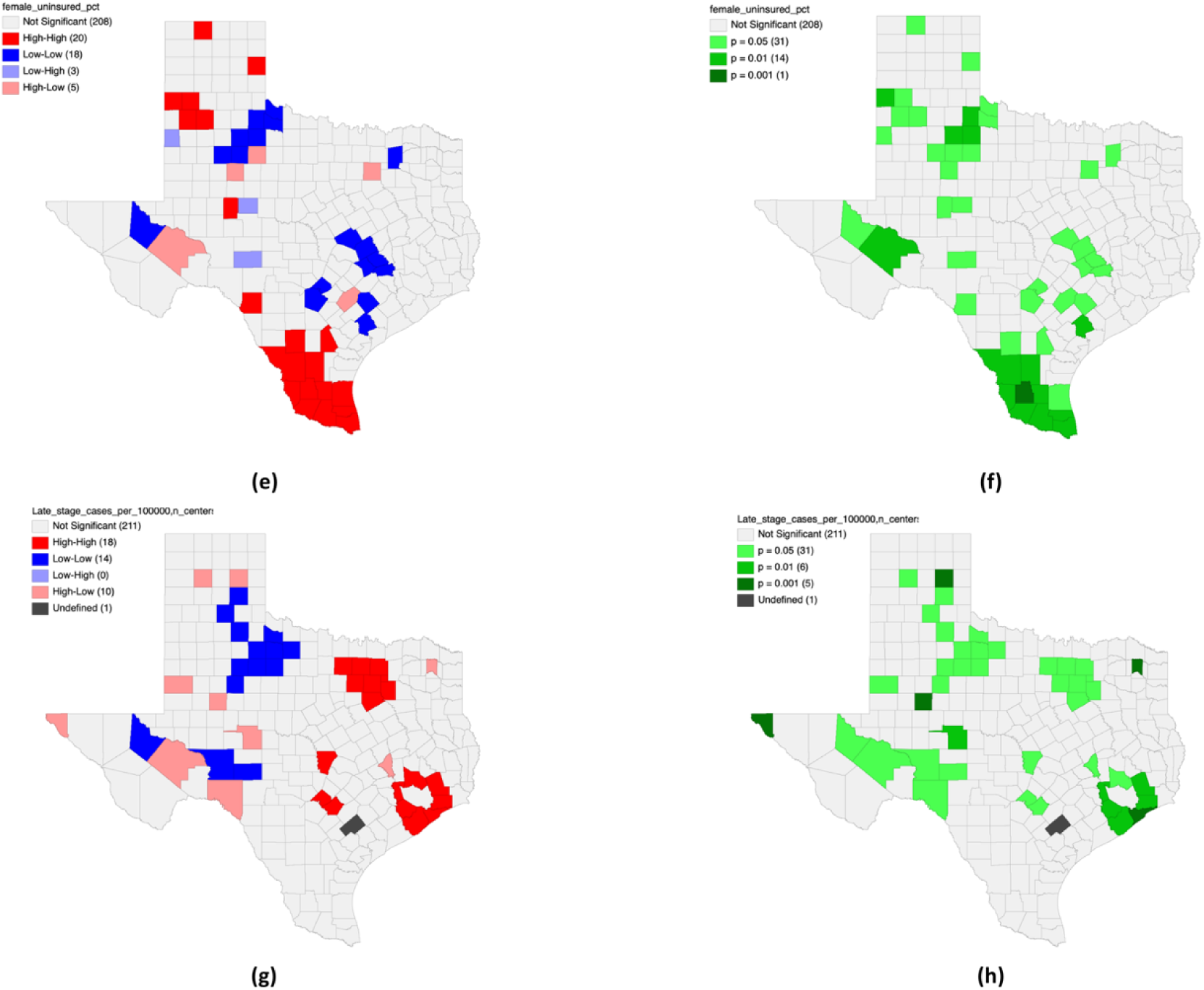
Univariate LISA for Uninsured Populations and Bivariate LISA for Incidence vs. Mammography Centers. (e) Univariate LISA cluster map for the percentage of uninsured females. (f) Corresponding univariate LISA significance map for uninsured females. (g) Bivariate LISA cluster map illustrating the local spatial correlation between late-stage incidence rates and the number of mammography centers. (h) Corresponding bivariate LISA significance map for late-stage incidence rates and mammography centers.

**Figure 5.**
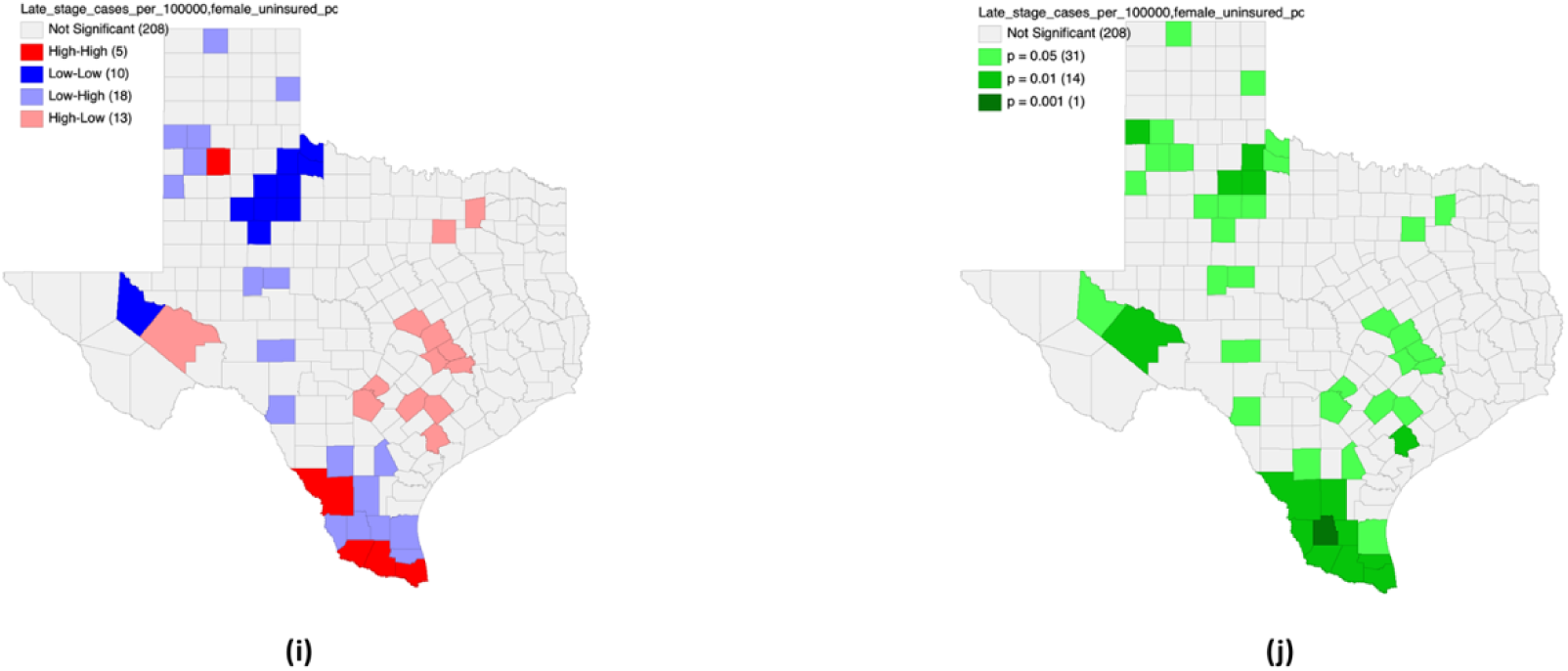
Bivariate LISA for Late-Stage Incidence vs. Uninsured Populations. (i) Bivariate LISA cluster map illustrating the local spatial correlation between late-stage incidence rates and the percentage of uninsured females. (j) Corresponding bivariate LISA significance map for late-stage incidence rates and the percentage of uninsured females.

### Bayesian Spatial Regression Analysis of Mammography Access, Urbanicity, and Late-Stage Breast Cancer Burden

In Table 2, Bayesian spatial Poisson conditional autoregressive regression demonstrated persistent geographic clustering in county-level late-stage breast cancer burden across Texas counties (ρ = 0.472, 95% CrI [0.038, 0.919]). After adjustment for female socioeconomic characteristics and residual spatial dependence, mammography centers per 100,000 female population demonstrated a borderline inverse association with late-stage breast cancer burden (β = -0.008, 95% CrI [-0.017, 0.002]), suggesting that greater screening infrastructure availability may contribute to lower late-stage breast cancer burden across Texas counties. Additionally, log-transformed population density demonstrated a significant inverse association with late-stage breast cancer burden (β = -0.136, 95% CrI [-0.175, -0.103]), indicating that less densely populated counties experienced substantially greater burden relative to more urbanized counties.

Educational vulnerability, measured as the percentage of adults without a high school diploma, also demonstrated a statistically significant association with late-stage breast cancer burden (β = -0.012, 95% CrI [-0.022, -0.001]). This finding should be interpreted cautiously because ecological county-level associations may reflect broader regional differences in healthcare utilization, healthcare infrastructure, population distribution, and diagnostic intensity rather than individual-level protective effects. Female unemployment percentage, female uninsured percentage, and female population composition were not independently associated with late-stage breast cancer burden after adjustment.

Model fit statistics (Table 3) indicated strong Bayesian model performance (DIC = 1172.71; WAIC = 1169.57), supporting the appropriateness of spatial disease mapping approaches for examining geographically clustered breast cancer disparities across Texas counties.

**Table 2.** Bayesian Spatial Poisson CAR Regression Examining Educational and Language Vulnerability Associated with Late-Stage Breast Cancer Burden Across Texas Counties.

| Variable | Posterior Mean ($\beta$) | 95% Credible Interval |
| --- | --- | --- |
| Mammography centers per 100,000 female population | -0.008* | [-0.017, 0.002] |
| Adults without high school diploma (%) | -0.012** | [-0.022, -0.001] |
| Log-transformed population density | -0.136*** | [-0.175, -0.103] |
| Female population | 0.000 | [0.000, 0.000] |
| Female population percentage (%) | -0.398 | [-2.360, 1.656] |
| Female unemployment percentage (%) | -1.450 | [-3.525, 0.648] |
| Female uninsured percentage (%) | 0.387 | [-0.942, 1.696] |
| Spatial autocorrelation parameter ($\rho$) | 0.472*** | [0.038, 0.919] |

**Note:** Bayesian spatial Poisson conditional autoregressive (CAR) regression models were estimated using county-level late-stage breast cancer burden across Texas counties. Positive coefficients indicate a greater late-stage breast cancer burden associated with increasing predictor values. Negative coefficients indicate a lower burden associated with increasing predictor values. CrI = credible interval.

**Table 3.** Model Fit Statistics.

| Statistic | Value |
| --- | --- |
| DIC | 1172.71 |
| WAIC | 1169.57 |
| LMPL | -588.38 |
| Posterior samples retained | 8,000 |
| Burn-in iterations | 20,000 |
| Total MCMC samples | 100,000 |

**Note:** DIC = Deviance Information Criterion; WAIC = Widely Applicable Information Criterion; LMPL = Log Marginal Predictive Likelihood. Lower DIC and WAIC values indicate improved model fit. \*\* indicates that the 95% credible interval excluded 0 with moderate posterior certainty. \*\*\* indicates that the 95% credible interval excluded 0 with strong posterior certainty.

## Discussion

The findings of this study demonstrated substantial geographic disparities in late-stage breast cancer burden, mammography access, and socioeconomic vulnerability across Texas counties. Spatial analyses revealed that mammography services were disproportionately concentrated in major metropolitan regions, while many rural and sparsely populated counties had comparatively limited screening infrastructure, which is consistent with other studies [11, 31–34]. Similarly, county-level late-stage breast cancer burden demonstrated significant geographic clustering, particularly across portions of East Texas, Central Texas, and selected southeastern counties. The geographic clustering patterns observed in parts of East Texas and the Gulf Coast may also reflect broader environmental and industrial risk contexts beyond healthcare accessibility alone. Several counties within the Houston-Baytown metropolitan and petrochemical corridor contain high concentrations of oil refining, chemical manufacturing, and industrial activity, which have historically raised concerns about exposure to environmental carcinogens and cumulative health risks [35–39]. Although environmental and occupational exposure variables were not directly measured in the current analysis, these place-based industrial and environmental contexts may contribute to the elevated cancer burden observed in certain regions and warrant further investigation in future spatial epidemiologic research. These findings suggest that geographic disparities in healthcare infrastructure and urbanicity may contribute substantially to differences in breast cancer outcomes across Texas.

The Bayesian spatial regression analyses further indicated that population density was significantly associated with late-stage breast cancer burden after accounting for spatial dependence and female socioeconomic characteristics. Less densely populated counties experienced greater late-stage breast cancer burden relative to more urbanized counties, suggesting that rurality and reduced healthcare infrastructure availability may contribute to delayed detection and poorer breast cancer outcomes [40–45]. Mammography center density additionally demonstrated a borderline inverse association with late-stage burden, indicating that greater screening infrastructure availability may potentially contribute to reduced burden, although the association did not fully exclude the null value within the credible interval.

The findings also suggest that healthcare infrastructure alone may not completely explain geographic disparities in late-stage breast cancer burden. Even in some metropolitan regions with relatively high mammography center availability, elevated late-stage burden persisted. These patterns indicate that additional structural barriers, including healthcare utilization, transportation limitations, healthcare affordability, delayed screening participation, and regional socioeconomic inequalities, may continue to influence breast cancer outcomes despite greater screening infrastructure availability.

From a public health and health services perspective, the findings support the need for geographically targeted breast cancer prevention and screening interventions across Texas. Rural and sparsely populated counties may benefit from expanded mammography infrastructure, mobile screening programs, transportation support services, and improved regional healthcare accessibility. Simultaneously, urban areas with persistent late-stage burden may require interventions focused on healthcare utilization, screening adherence, and reduction of socioeconomic barriers to preventive care. Collectively, the results demonstrate that breast cancer disparities across Texas are shaped by the intersection of geographic healthcare access, urbanicity, and broader structural determinants of health.

Several limitations should be considered when interpreting these findings. First, this study employed an ecological county-level design; therefore, associations identified at the county level should not be interpreted as individual-level relationships. Second, several counties contained suppressed or unavailable breast cancer data because of privacy protection procedures associated with small case counts, particularly in rural western Texas and Panhandle regions. These suppressed values may partially obscure localized spatial patterns. Third, the analyses relied on cross-sectional spatial relationships and therefore cannot establish causal relationships between healthcare infrastructure, socioeconomic characteristics, and breast cancer burden. Fourth, mammography center density does not directly measure screening utilization, healthcare quality, appointment availability, or transportation accessibility within counties. Finally, unmeasured contextual factors, including healthcare-seeking behaviors, physician availability, regional differences in healthcare systems, and racial or ethnic disparities, may also contribute to the observed geographic patterns.

## 5. Conclusion

This study demonstrated substantial geographic disparities in late-stage breast cancer burden across Texas counties and identified significant spatial clustering in mammography access and socioeconomic vulnerability. Bayesian spatial regression analyses indicated that lower population density was associated with greater late-stage breast cancer burden, highlighting the importance of urbanicity and healthcare infrastructure availability in shaping breast cancer outcomes across Texas. Mammography center density additionally demonstrated a borderline inverse association with late-stage burden, suggesting that expanded screening infrastructure may potentially contribute to improved early detection outcomes.

Overall, the findings emphasize the importance of geographically targeted breast cancer prevention and screening strategies that account for regional healthcare infrastructure disparities and rural healthcare access limitations. Spatial epidemiologic approaches may provide valuable tools for identifying underserved areas and informing precision public health interventions designed to reduce geographic inequities in breast cancer outcomes.

## Data Availability

No data was generated by this study. The following existing data sources were used: 1) Texas Cancer Information mammography database 2) National Cancer Institute cancer statistics databases 3) Centers for Disease Control and Prevention Social Vulnerability Index. And 4) National Historical Geographic Information System and United States Census-derived datasets. Additional analytic code and processed spatial datasets are available from the corresponding author upon reasonable request.

https://www.texascancer.info/prevscrntreat/mammography.html

https://www.statecancerprofiles.cancer.gov/incidencerates/

https://www.atsdr.cdc.gov/place-health/php/svi/svi-data-documentation-download.html

https://www.nhgis.org/data

https://data.census.gov/

## List of Abbreviations

ACS: American Community Survey
CAR: Conditional Autoregressive
CDC: Centers for Disease Control and Prevention
CrI: Credible Interval
DIC: Deviance Information Criterion
GIS: Geographic Information Systems
LISA: Local Indicators of Spatial Association
LMPL: Log Marginal Predictive Likelihood
NCI: National Cancer Institute
SVI: Social Vulnerability Index
WAIC: Widely Applicable Information Criterion

## Declarations

### Clinical trial number

Not applicable.

### Ethical Approval and Consent to Participate

This study utilized publicly available, county-level secondary data obtained from the National Cancer Institute, Texas Cancer Information database, United States Census-derived datasets, and the Centers for Disease Control and Prevention Social Vulnerability Index. Because the study used publicly available aggregated county-level data without individual identifiers or direct participant interaction, institutional review board approval and informed consent were not required. All methods were carried out in accordance with relevant guidelines and regulations.

### Consent for Publication

Not applicable.

### Availability of Data and Materials

The datasets analyzed during the current study are publicly available from the following sources: 1) Texas Cancer Information mammography database; 2) National Cancer Institute cancer statistics databases; 3) Centers for Disease Control and Prevention Social Vulnerability Index. And 4) National Historical Geographic Information System and United States Census-derived datasets. Additional analytic code and processed spatial datasets are available from the corresponding author upon reasonable request.

### Authors’ Contributions

Yue Zhang contributed to data acquisition, geographic information system mapping, spatial analyses, statistical interpretation, and drafting the manuscript. Jingjing Gao conceived the study design, supervised the study, spatial analyses, statistical modeling, data interpretation, and manuscript revision. Jilin Tian contributed to spatial epidemiologic interpretation and manuscript revision. Gayla M. Ferguson contributed to the public health interpretation of the findings and critically revised the manuscript. Jason H. Windett contributed to the spatial analytic framework, interpretation of geographic disparities, and manuscript revision. All authors reviewed and approved the final manuscript.

## Acknowledgements

The authors acknowledge the Texas Cancer Information program, the National Cancer Institute, the Centers for Disease Control and Prevention, and the National Historical Geographic Information System for providing publicly accessible datasets used in this study. The authors additionally acknowledge the developers of ArcGIS Pro, GeoDa, and R statistical software for providing tools used in the spatial analyses.

